# Dementia Prediction in Older People through Topic-cued Spontaneous Conversation

**DOI:** 10.1101/2021.05.18.21257366

**Authors:** Tomasz M. Rutkowski, Masato S. Abe, Seiki Tokunaga, Mihoko Otake-Matsuura

## Abstract

An increase in dementia cases is producing significant medical and economic pressure in many communities. This growing problem calls for the application of AI-based technologies to support early diagnostics, and for subsequent non-pharmacological cognitive interventions and mental well-being monitoring. We present a practical application of a machine learning (ML) model in the domain known as ‘AI for social good’. In particular, we focus on early dementia onset prediction from speech patterns in natural conversation situations. This paper explains our model and study results of conversational speech pattern-based prognostication of mild dementia onset indicated by predictive Mini-Mental State Exam (MMSE) scores. Experiments with elderly subjects are conducted in natural conversation situations, with four members in each study group. We analyze the resulting four-party conversation speech transcripts within a natural language processing (NLP) deep learning framework to obtain conversation embedding. With a fully connected deep learning model, we use the conversation topic changing distances for subsequent MMSE score prediction. This pilot study is conducted with Japanese elderly subjects within a healthy group. The best median MMSE prediction errors are at the level of 0.167, with a median coefficient of determination equal to 0.330 and a mean absolute error of 0.909. The results presented are easily reproducible for other languages by swapping the language model in the proposed deep-learning conversation embedding approach.

## 1 Introduction

Dementia, especially aging-associated memory decline, is a very critical global challenge in this century. Approximately 50 million older adults live with a dementia spectrum of neurocognitive disorders, as reported in recent assessments by the World Health Organization (WHO), and there are nearly 10 million new cases every year. Social welfare and mental well-being support in aging societies is becoming overloaded due to increased longevity and progression of dementia cases that significantly affect healthcare costs (Livingston et al., 2017). The Cabinet Office in Japan annually publishes a report on the aging society to address the situation (Japanese Government, 2019). The United Nations Sustainable Development Goal #3 entitled “Good Health and Well-being” (United Nations, 2019) also highlights the need to address the phenomenon with a focus on healthy aging, and it fosters well-being for all at all ages. Modern approaches to dementia, and particularly the most severe occurrence of Alzheimer’s disease (AD), demand the improvement of personalized treatments, relying not only on traditional pharmacological interventions but also on lifestyle modifications (Bredesen, 2017), as well as cognitive preservation strategies (Czaja et al., 2018; Reinhart and Nguyen, 2019). Modern societies also await the development of dementia early-onset prediction and successful preventive actions, as broadly discussed in (Livingston et al., 2017). Many established pharmacological treatments and contemporary ‘beyond-a-pill’ and ‘digital-pharma’ therapeutical interventions require reliable biomarkers. A research focus on advanced applications of electroencephalography (EEG) imaging methods (Chen et al., 2006; Mcbride et al., 2015; Horvath et al., 2018; Rutkowski et al., 2018; Rutkowski et al., 2019; Rutkowski et al., 2019, 2020) often requires a more clinical-level environment for successful application.

Language and discourse communication are among the most advanced tools of human civilization, as they afford abstract thought formation and exchange, spanning from daily conversations to cultural growth (Hauser et al., 2002; Michel et al., 2011). Language disorders often elucidate cognitive impairments or neural atrophy, as written or spoken communication processing employs many brain regions (Monsch et al., 1992; Snowdon et al., 1996). Previous studies on the relationship between language and cognitive functions have validated the use of linguistic features, such as verbal fluency, lexicon, semantic associations, and temporal patterns in spontaneous speech, in distinguishing people with dementia from healthy individuals (Monsch et al., 1992; Snowdon et al., 1996; Troyer et al., 1998; Hoffmann et al., 2010; Laws et al., 2010; Szatloczki et al., 2015; Fraser et al., 2016; Aramaki et al., 2016). Considering the development of practical home use applications, several research groups have focused on predicting mild cognitive impairment (MCI) or mild dementia onset using machine learning methods trained on large-size language corpora (Hernández-Domínguez et al., 2018; Fraser et al., 2019; Asgari et al., 2017). Recently it has become more manageable to collect and analyze large-size language datasets, such as daily conversations (Stark et al., 2020; Power et al., 2020), due to the development of applications (Otake et al., 2009; Otake-Matsuura, 2018; Otake-Matsuura et al., 2021), acoustic feature processing approaches (Fu et al., 2020; d. la Fuente Garcia et al., 2020), and natural language processing (NLP) algorithms (Bojanowski et al., 2016; Joulin et al., 2016a,b).

We propose a machine-learning (ML) approach, relating to the domain of AI for the social good. Successful application shall allow for computerized discrimination of early MCI or mild dementia onset, defined by the Alzheimer’s Association as a Mini-Mental State Exam (MMSE) score of 20 ⩽ *MMSE* ⩽ 24, as thoroughly reviewed by Christa Maree Stephan et al. (2013). MMSE was initially developed to evaluate cognitive functions, including registration attention and calculation, recall, language, ability to follow simple commands, and orientation Tuijl et al. (2012). We hypothesize that taking part in a multiparty conversation requires attention and remembering to stay on a topic, intact language skills, and registration related to working memory. Thus, estimating the MMSE scores from conversational behavior patterns is a solid and challenging approach since it shall limit the human examiner subjectivity and errors.A self-reported working-memory decline causing subjective cognitive impairment (SCI) is one of the early signs employed in the medical community (Emrani et al., 2018), and it often manifests in modified linguistic patterns (Eyigoz et al., 2020).

The contribution of the project reported is twofold. First, we present a novel conversation path length analysis based on a modern natural language processing methodology converting whole conversational segments into multidimensional vectors representing topics. Next, we propose a deep neural network model to predict MMSE scores from older adults’ conversation topic patterns, supported by encouraging results confirmed on a dataset of 65 participants. The paper is organized as follows. The next section describes the data collection, experimental, and machine learning tools used in the project. Results and discussion follow, together with proposed future research directions.

## 2 Materials and Methods

In the initial processing stages, we apply a speech data collection paradigm (Otake-Matsuura, 2018; Otake-Matsuura et al., 2021), as explained in Section 2.1, together with natural language processing using a word-to-vector (word2vec) conversation embedding analysis method (Bojanowski et al., 2016; Joulin et al., 2016b,a) in Section 2.2. As the current project’s original contribution, in Section 2.3 we propose conversation topic path analysis for mild dementia onset elucidation. Finally, a description of a machine learning model for prediction of the Mini-Mental State Exam (MMSE) score from conversational speech utterances follows in Section 2.4. (An entire data processing diagram is also presented in supplementary material Figure **??**.)

### 2.1 Recording of Four-party Short Conversations

In the current project, we collected speech utterances from free and topic-cued spontaneous conversation samples lasting approximately 30 minutes each (Otake-Matsuura, 2018; Otake-Matsuura et al., 2021), using headset microphones limiting the capture of crosstalk among participants. (There was no need for subsequent sound separation or speaker identification as each recorded audio channel contained the voice of a single speaker.) The 65 elderly subjects (average age of 72.6 ± 3.2 years old; 35 females; 30 males) participated voluntarily in the study, and they all gave informed written consent. Each subject participated in 12 weekly informal four-party chat meetings, which resulted in about 13 recordings for each (due to technical difficulties, some meetings resulted in two records). In the current conversational project, each meeting’s topic was decided one week in advance, and the subjects were instructed to prepare and stay within the topic limits. The topic limitation allowed for separate analysis of each participant’s speech, although the other participants’ influence was potentially confounding. The participants were instructed to make balanced contributions.

The participant groups were formed at random, and the subjects did not know each other, but in the course of 12 weekly meetings, they became familiar with one another. The study was reviewed and approved by the RIKEN Ethics Committee in Wako-shi, Saitama, Japan. The older Japanese adults were recruited from the aged people support focused Tokyo Silver Human Resources Center. We set exclusion criteria to limit participants to healthy people only as follows: no prior dementia diagnosis; no history of neurological disease; no medication known to affect the central nervous system; and no fewer than 24 points on the Japanese Mini-Mental State Examination (MMSE) (Sugishita et al., 2018). The assessment we conducted relied on medical interviews, neuropsychological tests (including the Digit Symbol Substitution Test (DSST) and Verbal Fluency Test (VFT)) and self-reports. The resulting participant MMSE scores were in the range of 24 − 30 (28.0 ± 1.46; mean ± standard deviation; detailed subject MMSE score distributions and the number of recorded conversations for each score are depicted in supplementary material Figures **??** and **??**, respectively).

### 2.2 Recorded Conversation Preprocessing

We first identified the audio recordings for each speaker and transcribed them for further machine learning-based processing using the Google Cloud Speech-to-Text service (Google, Mountain View, CA, 2018). The speech-to-text transcripts were then manually corrected by hired independent native Japanese speakers. In a second step, we automatically decomposed all the transcripts into separate word sequences and performed lemmatization using the MeCab library (ver. 0.996) (Kudo, 2006) in Python, ver. 3.8.4, which is a valuable method for Japanese language transcript morphological analysis utilizing conditional random fields (Kudo et al., 2004). Finally, we obtained formatted transcripts for each participant’s spoken passages segmented into words in all sessions. We next transformed the formatted text transcripts to vectors through a word2vec approach (Joulin et al., 2016b) using a pre-trained neural network model for the Japanese language (available thanks to Grave et al. (2018) and Mikolov et al. (2018) under a Creative Commons Attribution-ShareAlike License 3.0) to reconstruct linguistic contexts of words or whole sentences/expressions. Such a neural network takes as its input a large corpus of words, and it produces a resulting vector space, usually of several hundred dimensions-each unique word in the corpus denoted by the corresponding unique vector in the space (Bojanowski et al., 2016). Word vectors sharing familiar contexts in the corpus are located close to one another in the space (Joulin et al., 2016b). Word2vec is an especially computationally-efficient predictive model for discerning word embeddings from raw text. FastText is a valid word embedding method that is an extension of the word2vec model implemented in Python, ver. 3.8.4 (fastText, 2020). Instead of deriving vectors for words directly, fastText represents each word as an *n* − *gram* of characters (Bojanowski et al., 2016; Joulin et al., 2016b,a). Additionally, fastText works well with rare words, so even if a word is not present in the corpus, during training it can be broken down into *n* − *grams* to create its embeddings. We computed vector representations of entire sentences/expressions by a speaker in the conversation, as introduced by Joulin et al. (2016b). In the project presented, we used fastText pre-trained Japanese vectors (fastText, 2020). We decided to use a publicly available model for possible simple reproducibility of the proposed approach in the 157 available languages (Grave et al., 2018; Mikolov et al., 2018). We generated the embeddings using the original 300–dimensional vectors and the reduced versions to 16, 32, 64, and 100 dimensions, using fastText tools; Grave et al. (2018); Mikolov et al. (2018), which apply principal component analysis for dimensionality reduction. We present a comparison of the embedding vectors in the results section. For each four-party conversational event, we aggregated the topic feature matrices for each speaker separately. In each feature matrix, the row number indexed the speaker’s sentences/expressions, which we defined as an activity score in the conversation. The feature matrices thus created were subsequently used for topic path analysis and MMSE prediction, as discussed in the following sections.

### 2.3 Conversation Topic Switching Path Analysis

The sentences/expressions denoted by index *m* were transformed into embedding vectors *v*_*m,c,u*_ for conversations *c* and user (speaker) numbers *u* ∈ {1, 2, …, 65}, as explained in the previous section. Using fastText, pre-trained Japanese vectors, we next analyzed topic variability within each four-party conversational event.

#### 2.3.1 Whole Conversation Topic Switching Path Length

For each user *u* in a conversation meeting denoted by *c*, we calculated topic switching distances at time step *m*, using the Euclidean distance

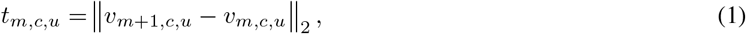

between two consecutive topic-representing vectors in a multidimensional word2vec-space and for *m* ∈ {1, …, 273 − 1} in the current project. We introduced a whole conversation topic switching path length *l*_*c,u*_ defined for a conversation number *c*, user number *u* as the sum of all progressive distances between topics mentioned (switched between) by each user separately in a single meeting, as

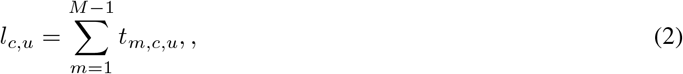

where *M* is the maximum number of the participant’s *u* conversation steps in a discourse *c*. The whole conversation topic switching path lengths thus obtained represent the number of different topics covered by each speaker.

#### 2.3.2 Speaker Activity in the Conversation

We calculated speaker activity *a*_*c,u*_ in a conversation *c* for each user *u* as the sum of all separate spoken contributions (discourse turns taken) in conversations represented by the multidimensional word2vec feature space in the project presented. In practice, the activity was the number *m* of word2vecs *v*_*m,c,u*_ for each participant in the conversation. Thus, the number of discourse contribution steps defined speaker activity in a single four-party conversation. We also compared pairwise the topic path lengths with the available user characteristics, namely age, gender, MMSE, and activity (number of turns by each speaker in each conversation), with results as explained in Section 3.1.

#### 2.3.3 Statistical Significance and Wasserstein Distances of Topic Distance Distributions between Pairs of Groups with the Same MMSE

We compared pairwise topic switching distance *t*_*m,c,u*_ distributions obtained from equation (1) at every conversation step *m* = 1 *∼*273 between pairs of groups of subjects with the same MMSE score. In order to do so, we first concatenated the results from all users *u* with the same MMSE score *s* and at every time step *m* into score-based topic distance continuous distribution functions *U*_*m,s*_ with *s ∈*{24, …, 30}. Next, we conducted statistical significance analyses using the Wilcoxon test and Wasserstein distance (Ramdas et al., 2017) (using NumPy by Harris et al. (2020) and SciPy by Virtanen et al. (2020) libraries in Python, ver. 3.8.4), comparing available topic distance distributions *U*_*m,s*_ for all pairs of available MMSE scores *s* at every time step *m*. We calculated Wilcoxon pairwise comparisons p-values for all pairs *i ≠ j* as *p*_*r,i,j*_(*m*) = ranksums (*U*_*m,i*_, *U*_*m,j*_), and Wasserstein distances *d*_*w,i,j*_(*m*) = inf (E[|*U*_*m,i*_, *U*_*m,j*_|]), where inf() is an infimum taken over all joint distributions of the random variables *U*_*m,i*_ and *U*_*m,j*_. *E*[*U*] denotes the expected value. The results of the two analyses above are discussed in Section 3.1.

### 2.4 MMSE Prediction from Small Group Conversation Patterns

We have proposed a simple yet powerful, fully connected neural network model with a linear output layer (Chollet, 2017) to predict MMSE-scores from conversational patterns focusing on spoken expression topic variability within a four-party discourse for each speaker separately. Often in real-life situations, we have to deal with randomness, and we do not always observe the same MMSE evaluations for similar cognitive performance by elderly subjects. A deep neural network model can learn to predict MMSE scores even from a limited number of training samples.

The challenging task with the fully connected neural network design is choosing the correct number of layers and learned parameters. The data type to be modeled plays a vital role in such a case. Although the cognitive MMSE scores are not continuous, we assume continuity, and we model the possible score values as a regression model. The fully connected neural network (FNN) model with two hidden layers employing rectified linear units (ReLU) was implemented in Keras by Chollet (2017) and TensorFlow by Abadi et al. (2015) within architecture as summarized in Table 1. (We also provide a full data processing diagram in supplementary material Figure **??**.) Four input features of the FNN models (as stated in Table 1) are composed of the whole conversation topic switching path length *l*_*c,u*_ obtained from equation (2), speaker activity *a*_*c,u*_, gender, and age. We trained the neural network with RMSprop optimizer (Goodfellow et al., 2016; Chollet, 2017) and a loss function of mean squared error. Training set batch sizes were set to 32 due to a limited training sample (844 in the study). Empirical training experiments with early stopping criterion suggested that 200 epochs would be sufficient for good validation results, avoiding overfitting. Therefore, we set the number for all ten-fold cross-validations (using a KFold function from the scikit-learn library by Pedregosa et al. (2011), with a shuffling of data before splitting into batches, and the same random state setting for all the tested dictionaries) and tested word2vec dictionary sizes for a fair comparison of the trained models. The KFold function guaranteed that the classifier training data from the same user’s conversation were not subsequently used to evaluate the same cross-validation run’s machine learning model. All the input features were normalized by subtracting the mean and dividing by the standard deviation.

**Table 1:** The fully connected neural network model applied in the study, implemented in Python, ver. 3.8.5, Keras library by Chollet (2017), and also in TensorFlow by Abadi et al. (2015)

| Layer type | Number of units | Activation function | Number of parameters |
| --- | --- | --- | --- |
| input layer | 4 | none | none |
| fully connected (dense) | 64 | ReLU | 320 |
| fully connected (dense) | 32 | ReLU | 2080 |
| fully connected (dense) | 1 | linear | 33 |

## 3 Results

The study results confirm the validity of the proposed approach to predict MMSE scores from conversational speech patterns focusing on topic switching. The new biomarker and machine learning model evaluation resulted in very encouraging outcomes supporting the original research hypothesis. We discuss the results in detail in the following Section 3.1 for conversation path analyzes and in 3.2 for MMSE predictions.

### 3.1 Conversation Topic Switching Path Analysis Results

The results of pairwise comparisons of the topic path lengths with available user characteristics, namely age, education, gender, MMSE, activity, DSST and VFT are summarized in Figure 1. The cognitive MMSE scores showed promising linear codependencies with topic path lengths, age, gender, and activity, which we explored next using a machine learning prediction approach, as discussed in the following section. Distributions of topic path lengths for participant groups with various MMSE scores are depicted in Figure 2, together with Wilcoxon rank-sums and Mann-Whitney U test significance scores. The group of 29 ≤ MMSE ≤ 30 produced statistically significantly longer path lengths at a Bonferroni-corrected significance level of *p* < 0.01667. MMSE ≤24 characterizes mild dementia onset (Christa Maree Stephan et al., 2013). Hence this preliminary result indicated that a more advanced deep neural network application was necessary. In Figure 3, we present statistical significances and Wasserstein distances (Ramdas et al., 2017). The analyses compared topic switching distributions during the discourses as a function of activity. The resulting statistical significance plots (depicted in the upper diagonal graphs in Figure 3) show conversation hot spots where the topic congruence varied. The more informative Wasserstein distances (presented in the lower diagonal graphs in the Figure 3) identify more clearly users from MMSE groups with scores of 24 and 25 as more gradually differing in the middle of conversations compared with the remaining pairwise evaluations. The two results above confirm the preliminary project hypothesis that the whole conversation topic switching path length was an appropriate input feature for subsequent machine learning model training.

**Figure 1:**
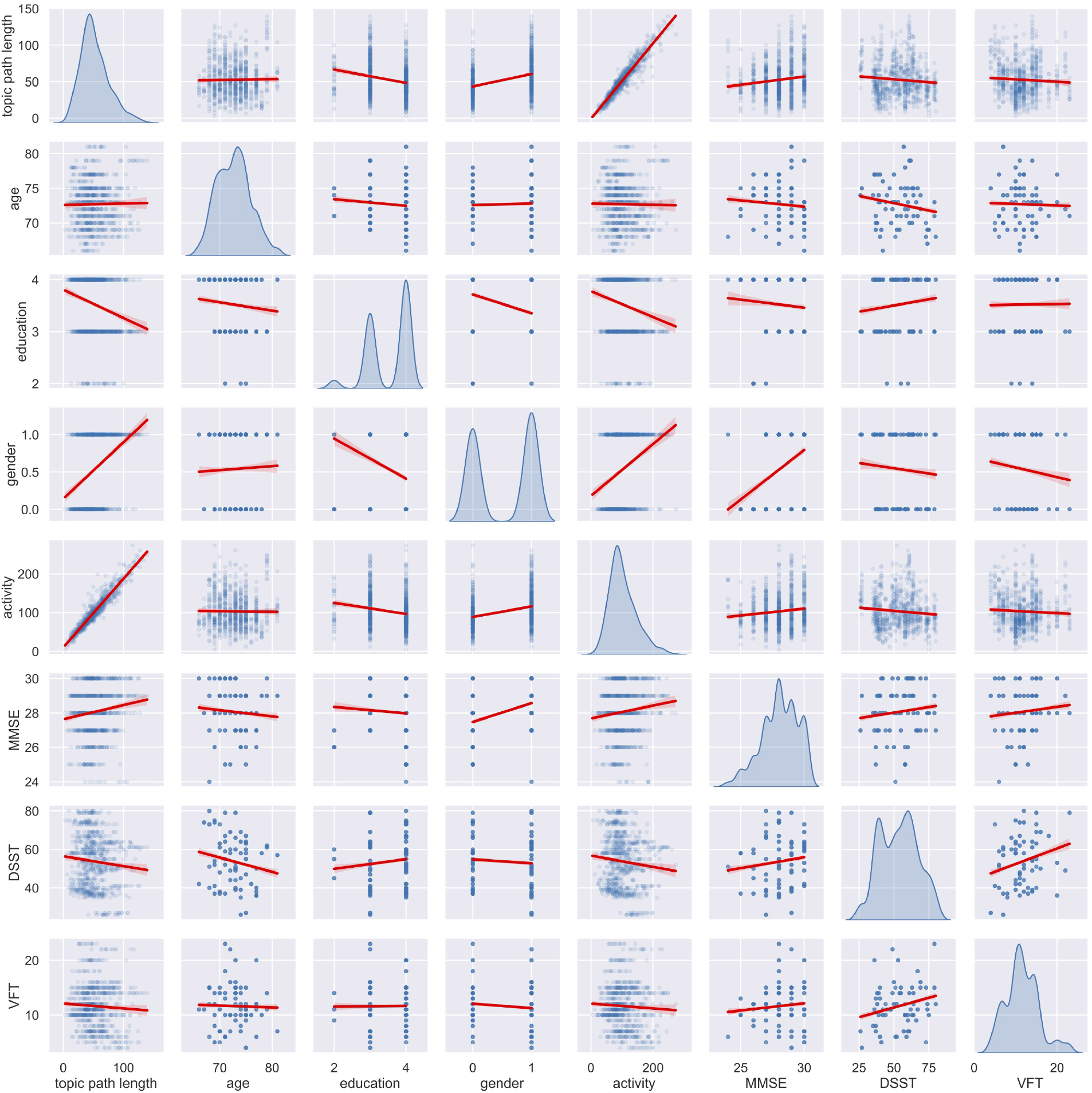
Pairwise plots show linear relations between all available feature pairs. In the gender feature category, females are denoted as 1, and males as 0. Education: indexed by 2 for 6 ∼ 9 years of education; 3 for 10 ∼ 12; 4 for 13 and above, respectively. DSST: Digit Symbol Substitution Test. VFT: Verbal Fluency Test. The results of the DSST and VFT display marginal correlations with MMSE, indicating no relation between topic path lengths and associative learning or semantic fluency, as estimated by these two tests. Within the project group, subject age, education, and gender also correlated only marginally with MMSE scores, VFT, DSST, and the proposed conversational features for the evaluated participants.

**Figure 2:**
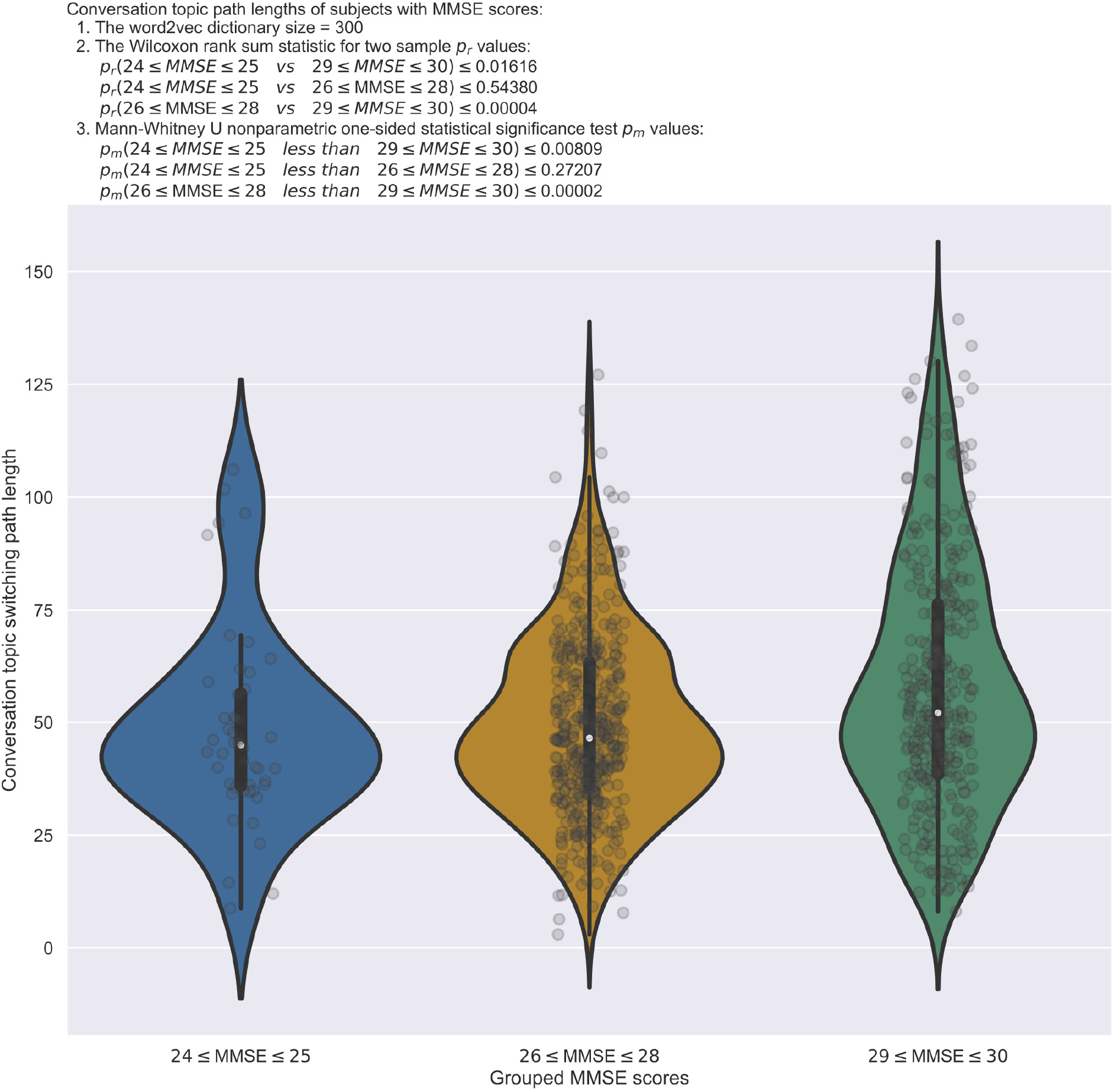
The violin plots depict the whole conversation topic switching path length distributions, obtained from equation (2) and binned into three groups of MMSE scores of the project’s subject population. The conversation topic path lengths of subject group 29 ≤ MMSE ≤ 30 are statistically significantly longer than evaluated with the Wilcoxon rank sums and Mann-Whitney U for larger median tests at a Bonferroni-corrected significance level of *p* < 0.01667.

**Figure 3:**
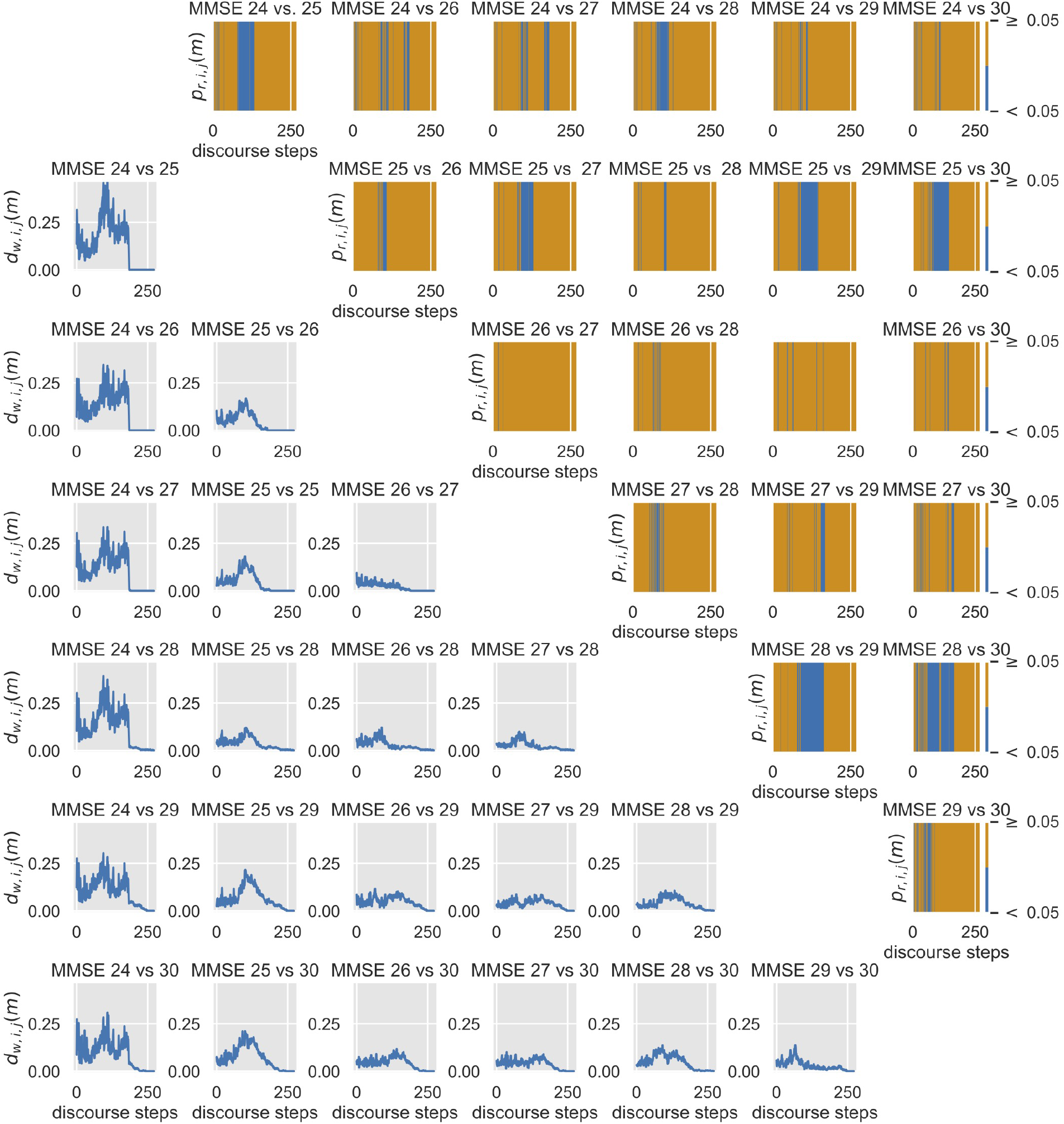
The upper diagonal graphs present pairs of distributions compared by means of Wilcoxon statistical significance tests during four-party conversations, with hot spots depicted in blue (*p*_*r,i,j*_(*m*) < 0.05) at every discourse contribution step *m*, where the topic congruence varied. In the lower diagonal traces with more informative Wasserstein distances *d*_*w,i,j*_(*m*) (Ramdas et al., 2017), pairwise evaluations identify more clearly speakers in MMSE groups with scores of 24 and 25 as more gradually differing in the middle of conversations compared with the remaining pairwise evaluations.

### 3.2 MMSE Prediction Results with Machine Learning Model

The encouraging ten-fold-cross-validation machine learning model results are summarized in the form of MMSE prediction error, coefficient of determination (*R*2), and mean absolute error (MAE) distributions for the word2vec dictionary sizes tested in Figure 4. The median predicted MMSE scores were similar for the all word2vec dictionaries tested with lower and upper quartiles in ranges of ± 1. The best median coefficients of determination (*R*2) and mean absolute errors are obtained for the most extensive dictionaries of sizes 100 and 300. The median results of all distributions depicted in Figure 4 are also summarized in Table 2 (scatter plots of real and predicted MMSE scores in supplementary materials Figure **??**). The sound predictions of MMSE based on conversational patterns confirm our project’s primary hypothesis of the possibility of estimating cognitive decline from speech patterns, focusing on topic switching paths in small group conversations.

**Table 2:** Median MMSE score prediction results using five different word2vec dictionary sizes

| Dictionary size of word2vec | Median MMSE error | Median $R^2$ | Median MAE |
| --- | --- | --- | --- |
| 16 | 0.457 | 0.248 | 0.988 |
| 32 | 0.520 | 0.231 | 0.985 |
| 64 | 0.337 | 0.265 | 0.953 |
| 100 | 0.199 | 0.303 | 0.917 |
| 300 | 0.167 | 0.330 | 0.909 |

**Figure 4:**
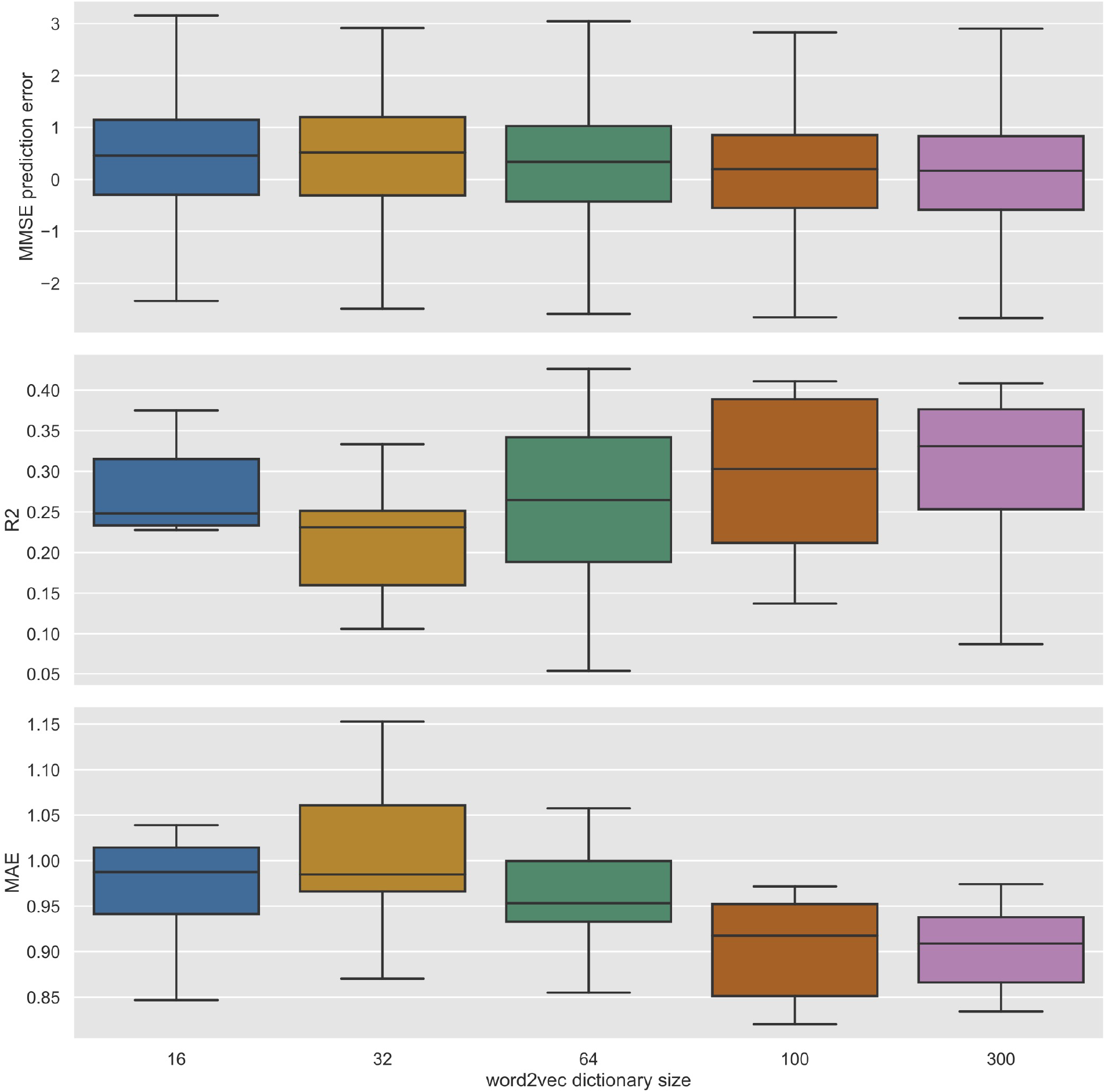
MMSE score predictions employing five different word2vec dictionary sizes using the proposed fully-connected deep neural networks. All predictions have mean errors below 1.0, and the best predictions for MMSE are for word2vec dictionary size 300.

## 4 Discussion

This feasibility study presents results obtained with a novel MMSE score prediction biomarker from conversational speech patterns. Although in the current project the MMSE score groups did not have the same number of subjects, the proposed fully connected neural network model produces encouraging results, as additionally explained in the scatter plot in supplementary materials Figure **??**. The growing success of machine learning methods contributing to modern-day AI applications has created many opportunities to employ recent natural language processing deep learning tools for social benefit. We have proposed a two-fold machine learning application. First, we employed publicly available speech-to-text and pre-trained fastText models for Japanese conversation vectorization in the so-called word2vec approach. The fastText pre-trained models are available for 157 languages; thus, our approach’s reproducibility is guaranteed. We show that the whole conversation topic switching path lengths obtained from each user discourse in the multidimensional space varied statistically for lower MMSE-scoring participants. These data were therefore suitable for the second machine learning model training. The final fully connected machine learning model, which is the present approach’s main contribution, allows for MMSE score prediction from recorded four-party conversation recordings. With smaller dictionaries, we also observed a reduction in computation complexity and time, and reduced memory requirements (the largest dictionary of size 300 was about 7*GB*, while the smallest of size 16 took 430*MB*). A future practical application in a smartphone or a wearable might benefit from a trade-off between accuracy and memory requirement.

The experimental results obtained in this study confirm the validity of the hypothesis of the possibility of employing machine learning methods to vectorize conversational utterances for subsequent topic switching analysis. The flattened word2vec features representing the conversation topics of each participant in a four-party conversation, with activity measures, age and gender used as input for the deep learning regressor training ten-fold cross-validation regime, are shown to result in the reliable prediction of participants’ MMSE scores. The encouraging median results offer a step forward in the development of novel methods, expected to improve the life of the elderly concerned about possible mental decline due to dementia. The potential employment of the aforementioned AI/ML-based mild dementia onset prediction may lead to a healthcare cost reduction, serving aging societies.

We also recognize the inherent deficiencies of the current method as our results are assessed on the basis of human-error-prone MMSE scores, which are only proxy estimators of cognitive decline. AI-based dementia predictors, if used without rigorous evaluation, might also pose a risk of misapplication or harm; therefore, stringent ethical measures would have to be implemented accordingly.

The preliminary results of simple path length analysis presented in Figure 2 produced mixed outcomes, showing that just a summation of topic distances was not adequate for a simple threshold-based application. In contrast, the application of a deep neural network model to the entire topic switching activity for each subject separately resulted in encouraging results for all word2vec dictionary sizes evaluated in the project, as shown in Figure 4 (also in the supplementary materials Figure **??**). A multiparty conversation evaluation could be implanted in teleconferencing systems, which have become very popular during the COVID-19 pandemic. We believe that MMSE score estimation within the reported limited prediction error could be acceptable as a biomarker component indicating possible cognitive decline from conversation patterns, without the necessity of older adults leaving their homes during pandemic lockdown situations. Pandemic lockdown-related isolation might accelerate dementia-related symptoms; therefore, simple conversation monitoring related measures could be implemented. Conversation topic-related word2vec features, which might be estimated on the subject’s mobile phone, would also guarantee privacy by not transmitting explicit discourse contents to dementia diagnostic centers.

In the next step of the project, we plan to evaluate the methods developed with a larger sample of healthy versus subjectively or mildly cognitively impaired individuals, and possibly as a biomarker monitoring tool for older adults diagnosed with dementia who are undergoing cognitive behavioral therapy. We furthermore intend to include a measure of daily function such as a clinical dementia rating scale (Morris, 1997), or a similar assessment to evaluate the older adult’s more broadly cognitive function. We also plan to combine the conversational measures with neurophysiological, especially EEG and fNIRS, methods for an even more trustworthy final objective evaluation of a possible cognitive decline due to age-related brain atrophy. We believe that the prospective involvement of AI techniques shall reduce or even partially reverse dementia pathologies in aging societies.

## Data Availability

The recorded speech data is not available due to privacy and ethical limitations.

## Author Contributions

TMR: Conceived the concept of the word2vec and conversation topic variability utilization for MMSE prediction with machine learning methods; MOM: designed and conducted conversational experiments with the elderly; ST: created and programmed the data collection environment; TMR created and programmed the data analysis; TMR and MSA: analyzed the data; TMR: wrote the paper.

## Acknowledgements

The company Handfast Point in The Netherlands proofread the manuscript. This research was supported in part by the KAKENHI, the Japan Society for the Promotion of Science Grant No. JP18K18140 (MSA), JP18KT0035 (MOM), JP19H01138 (MOM), JP20H05022 (MOM), JP20H05574 (MOM), and the Japan Science and Technology Agency AIP Trilateral AI Research Grant No. JPMJCR20G1 (MOM, TMR).

## A Supplementary Material

### A.1 Supplementary Figures

**Figure A.1:**
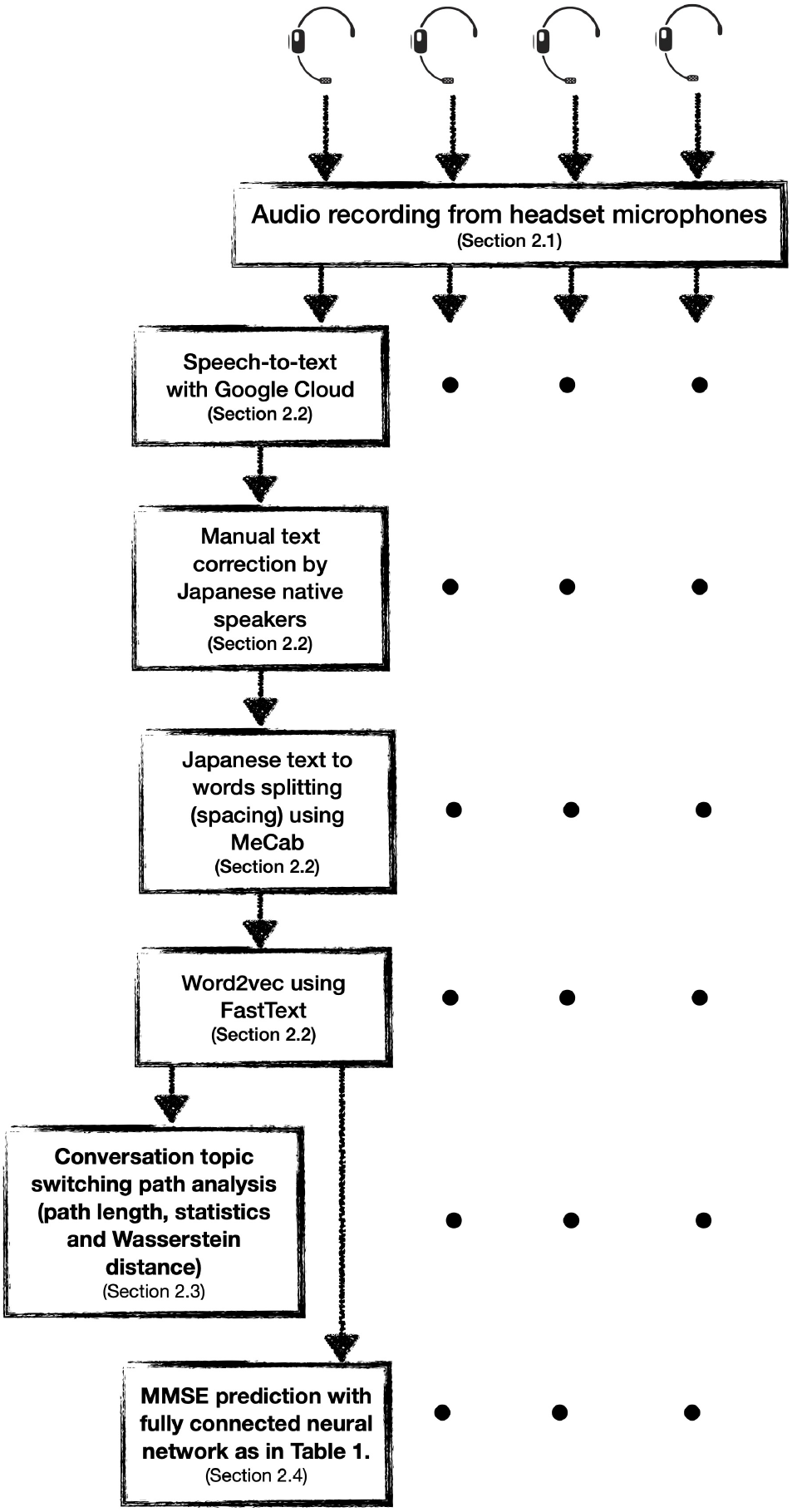
Full processing pipeline diagram from the recording of a four-party-conversation at the top to conversation topic switching analysis and MMSE prediction for each participant’s activity in a separate channel.

**Figure A.2:**
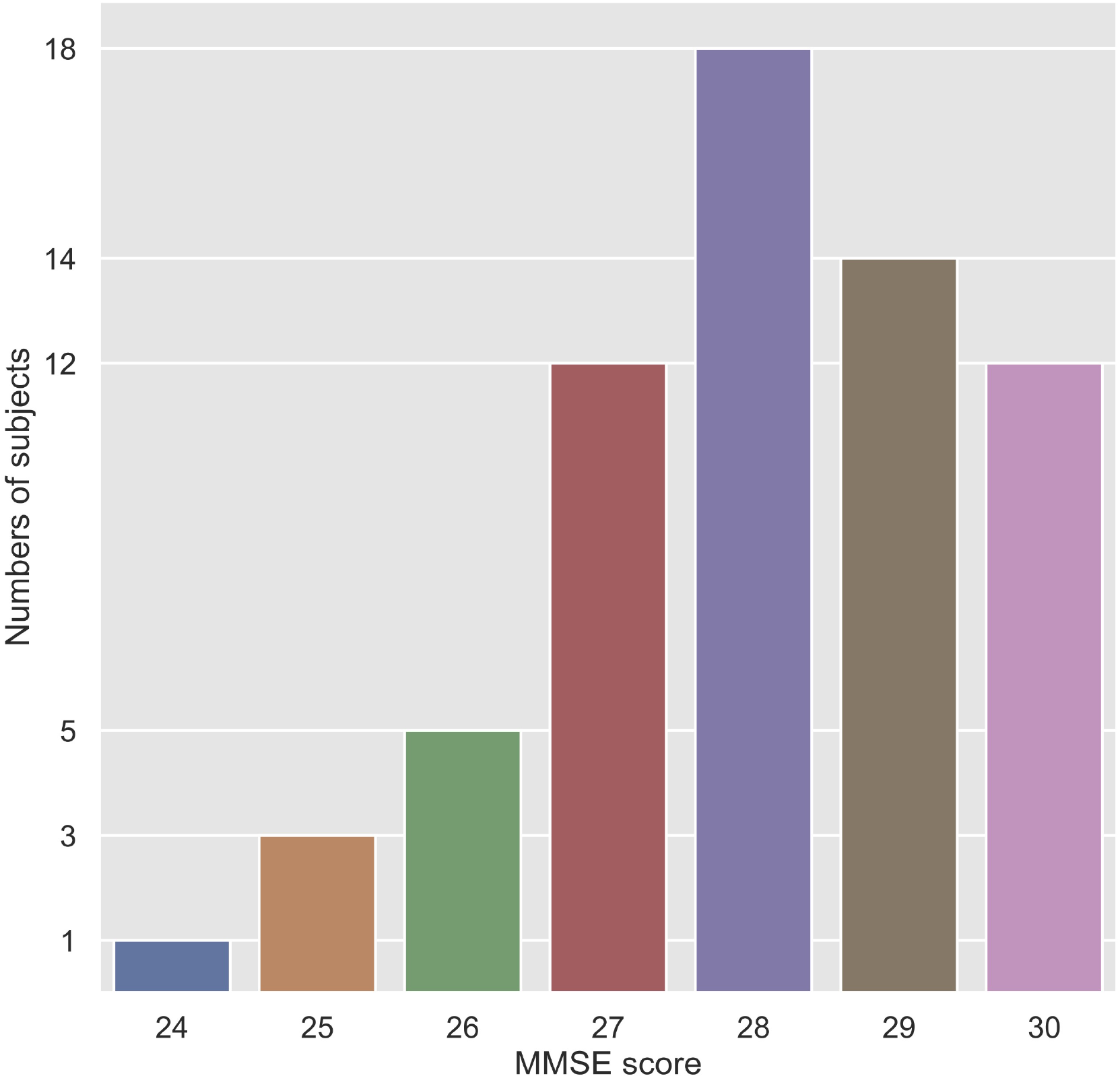
Numbers of subjects with each MMSE score in the dataset.

**Figure A.3:**
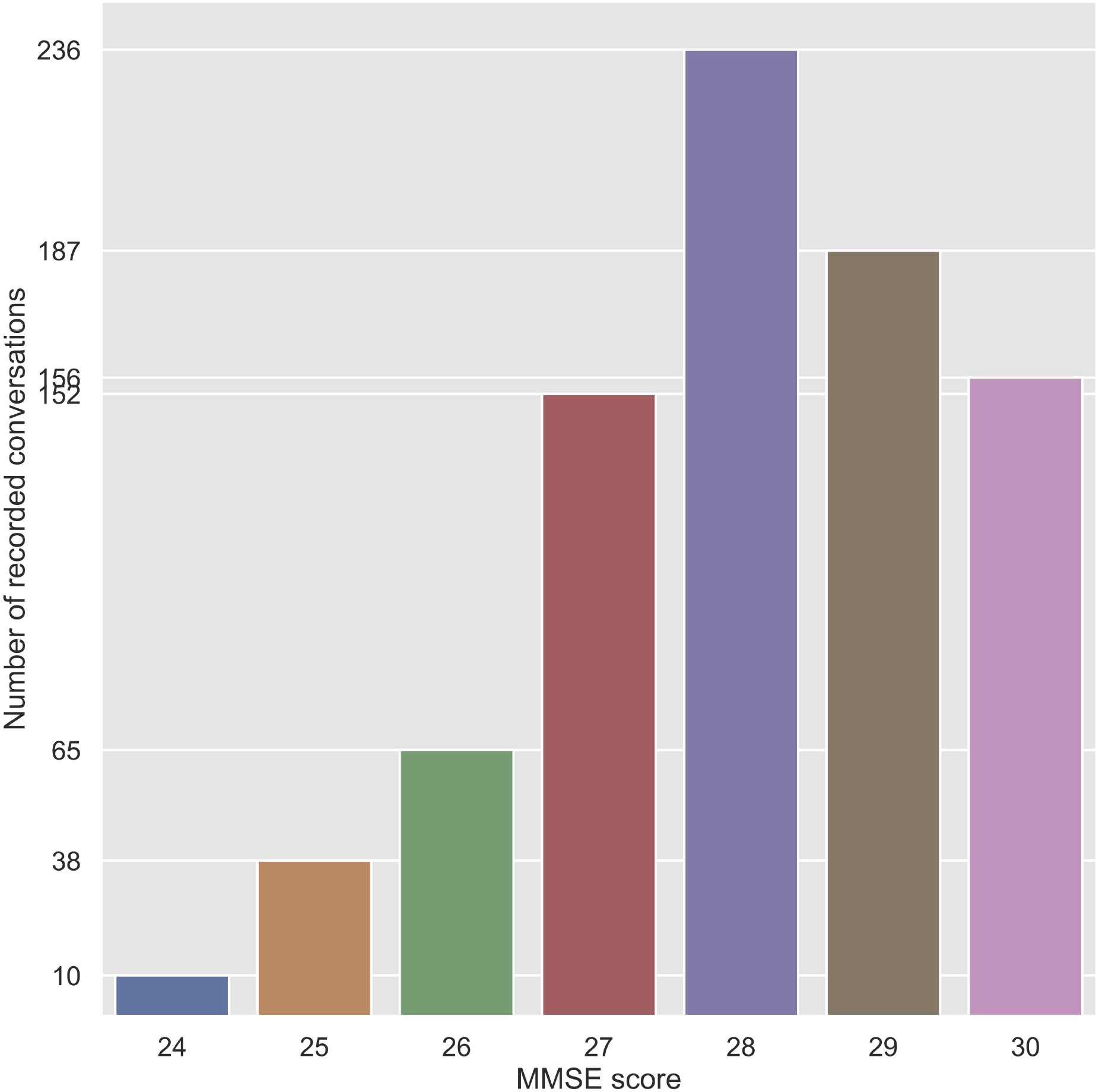
Number of recorded conversations for each MMSE score in the database. The numbers on the vertical axis add up to 844, which is the number of all the dataset samples. Each participant was intended to attend 12 conversational meetings, but some meetings were canceled, and some generated more than one recording.

**Figure A.4:**
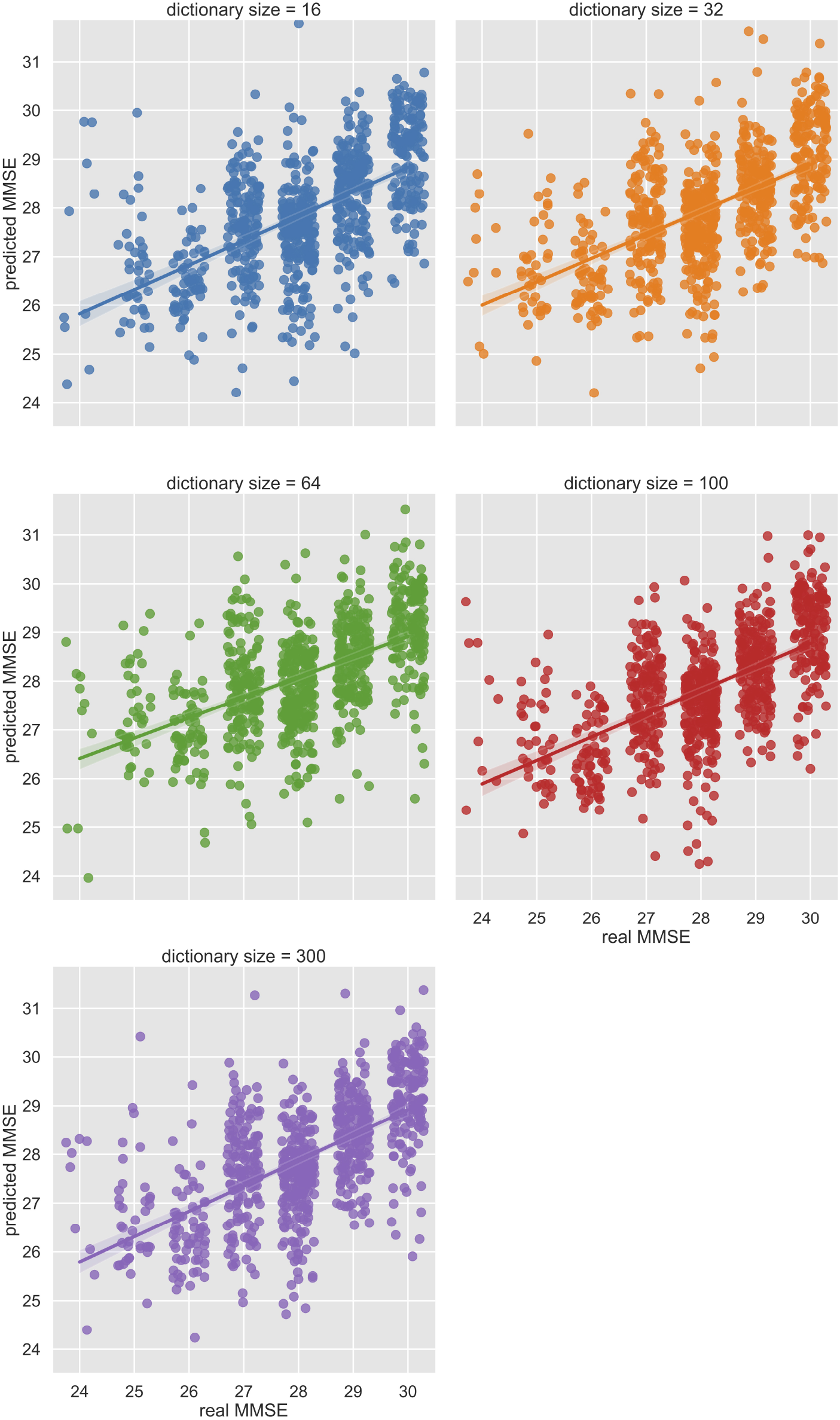
Scatter plots with real and predicted MMSE values using the proposed fully connected neural networks trained with word2vec dictionary sizes of 16, 32, 64, 100, and 300. The real MMSE values on the horizontal axis are randomly jittered in a range of ± 0.5 around the original values for visualization purposes. Figure created with Seaborn ver. 0.11.1 by Waskom and the seaborn development team (2020).

## Notes

### Competing Interest Statement

The authors have declared no competing interest.

### Clinical Trial

The study was reviewed and approved by the RIKEN Ethics Committee in Wako-shi, Saitama, Japan. RIKEN Center for Advanced Intelligence project, where the research was conducted, does not require clinical study registration. The study focuses on machine-learning-based biomarker prediction without a therapeutic application.

### Author Declarations

The study was reviewed and approved by the RIKEN Ethics Committee in Wako-shi, Saitama, Japan. The older Japanese adults were recruited from the aged people support focused Tokyo Silver Human Resources Center. The 65 elderly subjects participated voluntarily in the study, and they all gave informed written consent.

## References

Abadi, M., Agarwal, A., Barham, P., Brevdo, E., Chen, Z., Citro, C., et al. (2015). TensorFlow: Large-Scale Machine Learning on Heterogeneous Systems. Tech. rep. Software available from tensorflow.org

Aramaki, E., Shikata, S., Miyabe, M., and Kinoshita, A. (2016). Vocabulary size in speech may be an early indicator of cognitive impairment. PLOS ONE 11, e0155195. doi:10.1371/journal.pone.0155195

Asgari, M., Kaye, J., and Dodge, H. (2017). Predicting mild cognitive impairment from spontaneous spoken utterances. Alzheimer’s & Dementia: Translational Research & Clinical Interventions 3, 219–228

Bojanowski, P., Grave, E., Joulin, A., and Mikolov, T. (2016). Enriching word vectors with subword information. arXiv preprint 1607.04606

Bredesen, D. (2017). The End of Alzheimer’s: The First Programme to Prevent and Reverse the Cognitive Decline of Dementia (Vermilion)

Chen, Z., Cichocki, A., and Rutkowski, T. (2006). Constrained non-negative matrix factorization method for eeg analysis in early detection of alzheimer disease. In Acoustics, Speech and Signal Processing, 2006. ICASSP 2006 Proceedings. 2006 IEEE International Conference on. vol. V, 893–896. doi:10.1109/ICASSP.2006.1661420

[Dataset] Chollet, F. (2017). Deep learning with python

Christa Maree Stephan, B., Minett, T., Pagett, E., Siervo, M., Brayne, C., and McKeith, I. G. (2013). Diagnosing mild cognitive impairment (MCI) in clinical trials: A systematic review. BMJ Open 3, e001909. doi:10.1136/bmjopen-2012-001909

Czaja, S. J., Boot, W. R., Charness, N., Rogers, W. A., and Sharit, J. (2018). Improving Social Support for Older Adults Through Technology: Findings From the PRISM Randomized Controlled Trial. Gerontologist 58, 467–477. doi:10.1093/geront/gnw249

d. la Fuente Garcia, S., Haider, F., and Luz, S. (2020). Cross-corpus feature learning between spontaneous monologue and dialogue for automatic classification of alzheimer’s dementia speech. In 2020 42nd Annual International Conference of the IEEE Engineering in Medicine Biology Society (EMBC). 5851–5855. doi:10.1109/EMBC44109.2020.9176305

Emrani, S., Libon, D. J., Lamar, M., Price, C. C., Jefferson, A. L., Gifford, K. A., et al. (2018). Assessing working memory in mild cognitive impairment with serial order recall. J. Alzheimer’s Dis. 61, 917–928. doi:10.3233/JAD-170555

Eyigoz, E., Mathur, S., Cecchi, G., and Naylor, M. (2020). Linguistic markers predict onset of Alzheimer’s disease-NC-ND license (http://creativecommons.org/licenses/by-nc-nd/4.0/). EClinicalMedicine 0, 100583. doi:10.1016/j.eclinm.2020.100583

[Dataset] fastText (2020). fastText - library for efficient text classification and representation learning

Fraser, K. C., Fors, K. L., and Kokkinakis, D. (2019). Multilingual word embeddings for the assessment of narrative speech in mild cognitive impairment. Computer Speech & Language 53, 121–139

Fraser, K. C., Meltzer, J. A., and Rudzicz, F. (2016). Linguistic features identify alzheimer’s disease in narrative speech 49, 407–422. doi:10.3233/JAD-150520

Fu, Z., Haider, F., and Luz, S. (2020). Predicting mini-mental status examination scores through paralinguistic acoustic features of spontaneous speech. In 2020 42nd Annual International Conference of the IEEE Engineering in Medicine Biology Society (EMBC). 5548–5552. doi:10.1109/EMBC44109.2020.9175379

Goodfellow, I., Bengio, Y., and Courville, A. (2016). Deep Learning (MIT Press). http://www.deeplearningbook.org

Grave, E., Bojanowski, P., Gupta, P., Joulin, A., and Mikolov, T. (2018). Learning word vectors for 157 languages. In Proceedings of the International Conference on Language Resources and Evaluation (LREC 2018)

Harris, C. R., Millman, K. J., van der Walt, S. J., Gommers, R., Virtanen, P., Cournapeau, D., et al. (2020). Array programming with NumPy. Nature 585, 357–362. doi:10.1038/s41586-020-2649-2

Hauser, M. D., Chomsky, N., and Fitch, W. T. (2002). The faculty of language: what is it, who has it, and how did it evolve? science 298, 1569–1579

Hernández-Domínguez, L., Ratté, S., Sierra-Martínez, G., and Roche-Bergua, A. (2018). Computer-based evaluation of alzheimer’s disease and mild cognitive impairment patients during a picture description task. Alzheimer’s & Dementia: Diagnosis, Assessment & Disease Monitoring 10, 260–268

Hoffmann, I., Nemeth, D., Dye, C. D., Pákáski, M., Irinyi, T., and Kálmán, J. (2010). Temporal parameters of spontaneous speech in alzheimer’s disease. International Journal of Speech-Language Pathology 12, 29–34. doi:10.3109/17549500903137256. PMID: 20380247

Horvath, A., Szucs, A., Csukly, G., Sakovics, A., Stefanics, G., and Kamondi, A. (2018). EEG and ERP biomarkers of Alzheimer’s disease: a critical review. Frontiers in Bioscience 23, 183–220

Japanese Government (2019). Annual Report on the Aging Society. Tech. rep., Cabinet Office, Japan

Joulin, A., Grave, E., Bojanowski, P., Douze, M., Jégou, H., and Mikolov, T. (2016a). Fasttext.zip: Compressing text classification models. arXiv preprint 1612.03651

Joulin, A., Grave, E., Bojanowski, P., and Mikolov, T. (2016b). Bag of tricks for efficient text classification. arXiv preprint 1607.01759

Kudo, T. (2006). Mecab : Yet another part-of-speech and morphological analyzer. http://mecab.sourceforge.jp

Kudo, T., Yamamoto, K., and Matsumoto, Y. (2004). Applying conditional random fields to japanese morphological analysis. In Proceedings of the 2004 conference on empirical methods in natural language processing. 230–237

Laws, K. R., Duncan, A., and Gale, T. M. (2010). ‘normal’ semantic–phonemic fluency discrepancy in alzheimer’s disease? a meta-analytic study. Cortex 46, 595 –601. doi:https://doi.org/10.1016/j.cortex.2009.04.009

Livingston, G., Sommerlad, A., Orgeta, V., Costafreda, S. G., Huntley, J., Ames, D., et al. (2017). Dementia prevention, intervention, and care. The Lancet 390, 2673–2734

Mcbride, J., Zhao, X., Munro, N., Jicha, G., Smith, C., and Jiang, Y. (2015). Discrimination of Mild Cognitive Impairment and Alzheimer’s Disease Using Transfer Entropy Measures of Scalp EEG HHS Public Access. J Heal. Eng 6, 55–70. doi:10.1260/2040-2295.6.1.55

Michel, J.-B., Shen, Y. K., Aiden, A. P., Veres, A., Gray, M. K., Pickett, J. P., et al. (2011). Quantitative analysis of culture using millions of digitized books. science 331, 176–182

Mikolov, T., Grave, E., Bojanowski, P., Puhrsch, C., and Joulin, A. (2018). Advances in pre-training distributed word representations. In Proceedings of the International Conference on Language Resources and Evaluation (LREC 2018)

Monsch, A. U., Bondi, M. W., Butters, N., Salmon, D. P., Katzman, R., and Thal, L. J. (1992). Comparisons of verbal fluency tasks in the detection of dementia of the alzheimer type. Archives of neurology 49, 1253–1258

Morris, J. C. (1997). Clinical dementia rating: a reliable and valid diagnostic and staging measure for dementia of the Alzheimer type. International Psychogeriatrics 9, 173–176

Otake, M., Kato, M., Takagi, T., and Asama, H. (2009). Coimagination method: Communication support system with collected images and its evaluation via memory task. In International Conference on Universal Access in Human-Computer Interaction (Springer), 403–411

Otake-Matsuura, M. (2018). Conversation assistive technology for maintaining cognitive health. J Korean Gerontol Nurs 20, 154–159. doi:10.17079/jkgn.2018.20.s1.s154

Otake-Matsuura, M., Tokunaga, S., Watanabe, K., Abe, M. S., Sekiguchi, T., Sugimoto, H., et al. (2021). Cognitive intervention through photo-integrated conversation moderated by robots (PICMOR) program: A randomized controlled trial. Frontiers in Robotics and AI 8, 54. doi:10.3389/frobt.2021.633076

Pedregosa, F., Varoquaux, G., Gramfort, A., Michel, V., Thirion, B., Grisel, O., et al. (2011). Scikit-learn: Machine learning in Python. Journal of Machine Learning Research 12, 2825–2830

Power, E., Weir, S., Richardson, J., Fromm, D., Forbes, M., MacWhinney, B., et al. (2020). Patterns of narrative discourse in early recovery following severe traumatic brain injury. Brain Injury 34, 98–109. doi:10.1080/02699052.2019.1682192. PMID: 31661629

Ramdas, A., Trillos, N. G., and Cuturi, M. (2017). On wasserstein two-sample testing and related families of nonparametric tests. Entropy 19, 47

Reinhart, R. M. G. and Nguyen, J. A. (2019). Working memory revived in older adults by synchronizing rhythmic brain circuits. Nature Neuroscience 22, 820–827. doi:10.1038/s41593-019-0371-x

Rutkowski, T. M., Abe, M. S., Koculak, M., and Otake-Matsuura, M. (2019). Cognitive assessment estimation from behavioral responses in emotional faces evaluation task - ai regression approach for dementia onset prediction in aging societies -. In NeurIPS Joint Workshop on AI for Social Good (Vancouver, Canada), vol. Track 1 - Producing Good Outcomes, 1–4

Rutkowski, T. M., Abe, M. S., Koculak, M., and Otake-Matsuura, M. (2020). Classifying mild cognitive impairment from behavioral responses in emotional arousal and valence evaluation task - ai approach for early dementia biomarker in aging societies -. In The 42nd Annual International Conference of the IEEE Engineering in Medicine and Biology Society (EMBC). IEEE Engineering in Medicine and Biology Society (Montreal, Canada: IEEE Press), 5537–5543

Rutkowski, T. M., Koculak, M., Abe, M. S., and Otake-Matsuura, M. (2019). Brain correlates of task–load and dementia elucidation with tensor machine learning using oddball BCI paradigm. In ICASSP 2019 - 2019 IEEE International Conference on Acoustics, Speech and Signal Processing (ICASSP). 8578–8582. doi:10.1109/ICASSP.2019.8682387

Rutkowski, T. M., Zhao, Q., Abe, M. S., and Otake, M. (2018). AI neurotechnology for aging societies - task-load and dementia EEG digital biomarker development using information geometry machine learning methods. In AI for Social Good Workshop at the Neural Information Processing Systems (NeurIPS) 2018 (Montreal, Canada), 1–4

Snowdon, D. A., Kemper, S. J., Mortimer, J. A., Greiner, L. H., Wekstein, D. R., and Markesbery, W. R. (1996). Linguistic ability in early life and cognitive function and alzheimer’s disease in late life: Findings from the nun study. Jama 275, 528–532

Stark, B. C., Dutta, M., Murray, L. L., Bryant, L., Fromm, D., MacWhinney, B., et al. (2020). Standardizing assessment of spoken discourse in aphasia: A working group with deliverables. American Journal of Speech-Language Pathology 25

Sugishita, M., Koshizuka, Y., Sudou, S., Sugishita, K., Hemmi, I., Karasawa, H., et al. (2018). The validity and reliability of the Japanese version of the mini-mental state examination (mmse-j) with the original procedure of the attention and calculation task (2001). Ninchi Shinkei Kagaku 20, 91–110

Szatloczki, G., Hoffmann, I., Vincze, V., Kalman, J., and Pakaski, M. (2015). Speaking in Alzheimer’s disease, is that an early sign? Importance of changes in language abilities in Alzheimer’s disease. Frontiers in Aging Neuroscience 7, 195. doi:10.3389/fnagi.2015.00195

Troyer, A. K., Moscovitch, M., Winocur, G., Leach, L. Freedman, and Morris (1998). Clustering and switching on verbal fluency tests in alzheimer’s and parkinson’s disease. Journal of the International Neuropsychological Society 4, 137–143. doi:10.1017/S1355617798001374

Tuijl, J. P., Scholte, E. M., de Craen, A. J., and van der Mast, R. C. (2012). Screening for cognitive impairment in older general hospital patients: comparison of the six-item cognitive impairment test with the mini-mental state examination. International journal of geriatric psychiatry 27, 755–762

United Nations (2019). United Nations Sustainable Development Goals. Tech. rep.

Virtanen, P., Gommers, R., Oliphant, T. E., Haberland, M., Reddy, T., Cournapeau, D., et al. (2020). SciPy 1.0: Fundamental Algorithms for Scientific Computing in Python. Nature Methods 17, 261–272. doi:10.1038/s41592-019-0686-2

[Dataset] Waskom, M. tand the seaborn development team (2020). mwaskom/seaborn. doi:10.5281/zenodo.592845

